# Nicotinamide and Pyridoxine supplementation stimulates muscle stem cells in a randomized clinical trial on muscle repair

**DOI:** 10.1101/2025.03.24.25323226

**Authors:** Grith Højfeldt, Joris Michaud, Ann Damgaard, Karoline Karlog, Eugenia Migliavacca, Sonia Karaz, Elham P. Micol, Odd E. Johansen, Leonidas G. Karagounis, Bjørk W. Helge, William Hagemann, Michael Kjaer, Jerome N. Feige, Pascal Stuelsatz, Abigail L. Mackey

**Author notes:** These authors contributed equally (Grith Højfeldt, Joris Michaud). Shared senior authorship (Jerome N. Feige, Pascal Stuelsatz, Abigail L. Mackey).

## Abstract

Skeletal muscle regeneration is a cardinal feature of muscle pathologies and is crucial for post-exercise recovery and traumatic sports injuries. Regeneration of damaged muscle in humans is a prolonged process and is accompanied by pain and physical dysfunction, highlighting the unmet need for effective interventions to accelerate the regenerative process. Through cellular and preclinical models, we have previously identified nicotinamide (NAM) and pyridoxine (PN) as potent stimulators of Muscle Stem Cells (MuSCs). Herein we investigated if a combination of NAM and PN could enhance MuSC activity and improve muscle regeneration in healthy volunteers during recovery from eccentric contractions.

**Methods:** This randomized, double-blind, placebo-controlled trial enrolled male participants aged 18-50 years supplemented daily with 714mg NAM and 19mg PN (NAM/PN) or placebo for 8 days following unilateral eccentric muscle contractions using Neuromuscular Electrical Stimulation (NMES). MuSC was quantified by immunohistofluorescence on vastus lateralis muscle biopsies.

**Results:** 39 out of 43 enrolled participants completed the study. Supplementation of NAM/PN was well tolerated and increased blood concentrations of NAM and PN vitamers. The NMES protocol caused myofiber necrosis and triggered a strong MuSC response. After 8 days, the number of Pax7, MyoD, and myogenin positive cells per damaged fiber was significantly higher in NAM/PN vs placebo groups (+29-67%). NAM/PN also increased the proportion of regenerating fibers re-expressing embryonic myosin (+37%).

**Conclusion:** Daily oral NAM/PN supplementation following eccentric muscle damaging contractions enhances MuSC activity and accelerates muscle regeneration. These findings provide new possibilities for targeted therapeutic interventions in muscle repair.

**Trial registration:** NCT04874662

**One Sentence Summary:** Muscle regeneration is enhanced by nicotinamide and pyridoxine supplementation, accelerating recovery and offering therapeutic potential.

## INTRODUCTION

Muscle Stem Cells (MuSCs), skeletal muscle tissue resident stem cells, primarily drive muscle repair during regeneration after myofiber injury (*1*) but also contribute to muscle growth (*2, 3*) and long-term maintenance (*4*). Specifically, in addition to their capacity to ensure repair after overt trauma involving myofiber necrosis, MuSCs support the turnover of myonuclei during muscle homeostasis and fuse with myofibers to maintain myonuclear domains, facilitating resistance training-induced muscle hypertrophy (*5, 6*). Under normal conditions, MuSCs remain in a quiescent state. However, upon activation by signals from the niche, MuSCs proliferate, differentiate, and either fuse with existing myofibers or with each other to replace necrotic myofibers. They can also self-renew to replenish the MuSC pool. At the molecular level, MuSCs and their progeny are tightly regulated by sequential expression of transcription factors, notably Pax7 and the myogenic regulatory factors MyoD and myogenin, allowing for monitoring MuSC progression (*7*).

Maintaining healthy skeletal muscle is a major determinant of the quality of life across the lifespan and MuSCs play a crucial role in preserving its regenerative and adaptive capacity. Skeletal muscle injuries are common occurrences and are frequently observed in both recreational and professional sports as a result of strains and contusions (*8–11*). Strenuous and unaccustomed exercise, especially exercise involving eccentric muscle contractions where a muscle lengthens while generating force, are known to induce damage to the cellular structures and the extracellular matrix. This type of contraction is typically involved in exercises where the muscle is actively resisting the external load or gravity, such as weightlifting, running downhill, jumping, squatting or other strength training exercises like pull-ups or step-ups. Other causes of muscle injuries include laceration resulting from accidents during activities of daily living and in the workplace, or iatrogenic muscle damage following surgical procedures (*12*). While skeletal muscle possesses the ability to recover from tissue damage, the process of muscle regeneration is often slow, extending beyond four weeks (*13*), hindering the return to regular activities. Moreover, a range of conditions such as aging (*14*), cachexia (*15–17*), diabetes (*18, 19*), and muscular dystrophies (*20, 21*), disrupt myofiber integrity and impair the muscle’s natural ability to regenerate and repair. Despite the high prevalence of muscle injuries, traditional therapeutic options for treating damaged muscles are limited and include variations on RICE (Rest, Ice, Compression and Elevation) , POLICE (Protection, Optimal Loading, Ice, Compression and Elevation) (*22*) and more recently PEACE and LOVE (Protection, Elevation, Avoid Anti-inflammation, Compression, Education & Load, Optimism, Vascularisation and Exercise) protocols, followed by physical therapy and rehabilitation exercises(*23*). The use of nonsteroidal anti-inflammatory drugs (NSAIDs) shortly after a muscle injury can provide pain relief and reduce inflammation and swelling (*24*). However, there is conflicting data regarding their long-term effects on muscle regeneration. Some studies suggest that NSAIDs may offer little to no benefit in promoting muscle healing and may even have detrimental effects that delay the recovery process (*25*), and the current PEACE and LOVE guidelines advise against NSAID use. Importantly, there is a lack of therapeutic interventions that effectively support muscle recovery by directly targeting the endogenous regenerative potential of skeletal muscle.

In a previous study, we used a high-content imaging screen of a library of over 50,000 natural bioactive molecules and food-derived nutrients on human myogenic progenitors to identify novel nutritional molecules targeting MuSCs (*26*). Using this approach, we discovered that nicotinamide (NAM) and pyridoxine are potent nutrients that stimulate MuSC proliferation and induce their differentiation. We then tested the efficacy of oral NAM/PN supplementation in two different preclinical models of muscle regeneration, after an acute muscle injury induced with cardiotoxin and in a model of eccentric contraction-induced muscle regeneration. Based on our collective findings, NAM/PN has emerged as a promising nutritional intervention that stimulates MuSCs and enhances muscle repair, with potential for direct translation into clinical applications.

While preclinical models of muscle regeneration are well-established (*27*), studying muscle regeneration in humans is more complex as it requires a standardization of the complex physiological and molecular aspects that drive muscle repair and recovery. Human models of muscle recovery, typically exercise-based, often exhibit high inter-individual variability and may not always result in myofiber necrosis, indicating more of a muscle remodeling process rather than true regeneration. To overcome this limitation, we have developed a technique that combines neuromuscular electrical muscle stimulation (NMES) with forced lengthening contractions in a controlled setting (*28–32*). This approach induces myofiber necrosis followed by muscle regeneration with full restoration of force producing capacity and no long-term side effects.

Notably, while most other models succeed at triggering the initial phase of muscle regeneration as indicated by the expansion of Pax7^+^ MuSCs, a robust increase in differentiating progenies (myogenin^+^ cells and eMyHC^+^ fibers) has been observed primarily with this NMES protocol (*30, 31, 33–35*).

We hypothesized that the oral NAM/PN nutritional intervention, already established to enhance muscle repair in rodents, would also improve MuSC-mediated repair in a human model of muscle regeneration following NMES eccentric contractions, providing evidence for use in clinical settings. To this end, the pre-registered primary outcome was the number of MuSCs (Pax7 and myogenin) on sections of muscle biopsies from the stimulated leg, comparing the study intervention (NAM/PN) to the control intervention (placebo).

## RESULTS

The primary objective of this study was to investigate if a combination of nicotinamide (NAM) and pyridoxine (PN) intake would improve human skeletal muscle regeneration, through enhanced activation of MuSCs. To this end, the primary endpoint was the number of MuSCs measured by immunofluorescence on sections of muscle biopsies during experimentally-induced muscle regeneration. An overview of the study design and study flow diagram are presented in Figure 1A,B. Forty-three healthy males aged 18 to 50 years were enrolled and randomly assigned to supplementation with either NAM/PN (NAM: 714 mg/day, PN: 19 mg/day, n=22) or placebo (n=21) (Figure 1B, see Table 1 for an overview of participant characteristics). One participant prematurely withdrew, and three participants had a major protocol deviation independent of the treatment (two missed the last visit and one Day 8 muscle biopsy was not usable). Thus, 39 participants completed the study (19 NAM/PN, 20 placebo).

**Figure 1:**
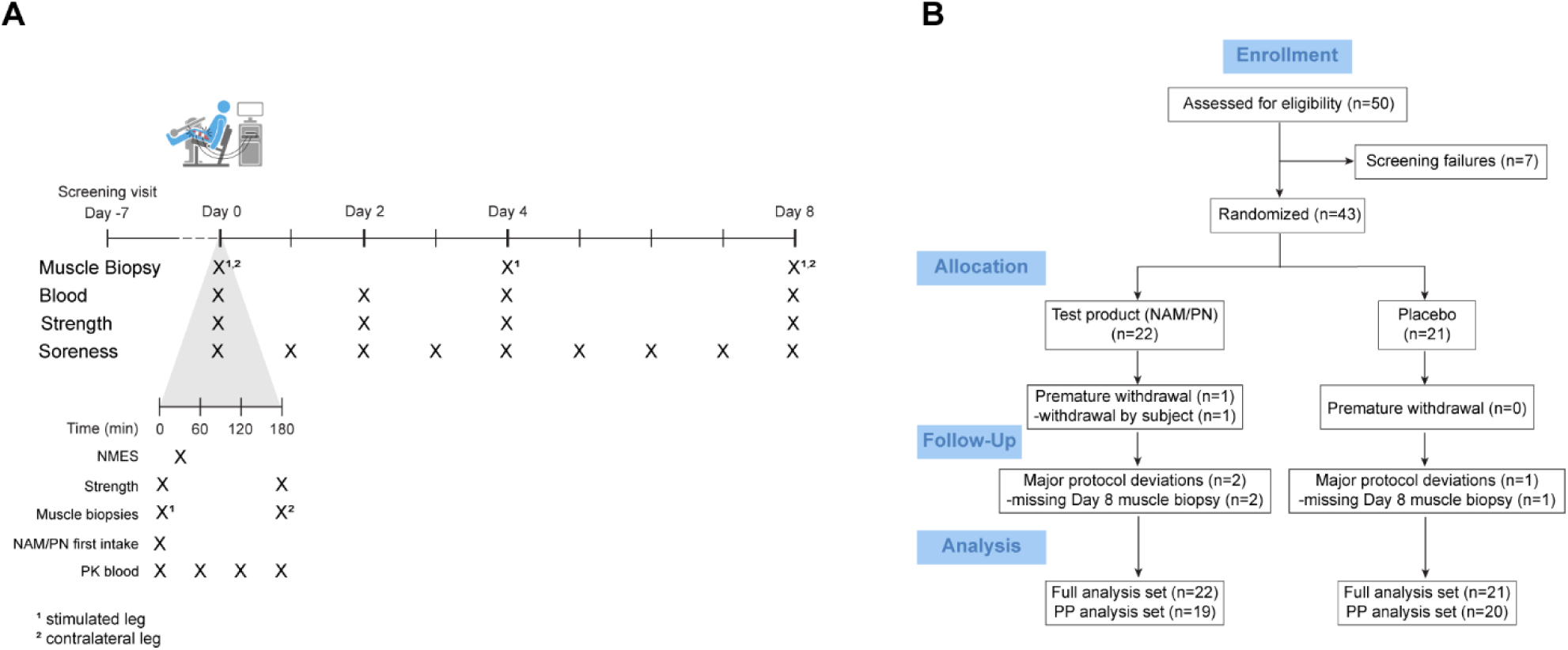
Study design and study flow diagram. (A) Overview of the study design indicating the timing of muscle biopsies, blood samplings, measurements of muscle strength and soreness during the 9 days period of NAM/PN or placebo intake in healthy male participants. The bottom timeline displays in more detail the timing and order of events during the 3h protocol at Day 0 when Neuromuscular electrical stimulation (NMES) was performed. (B) CONSORT flow chart.

**Table 1.**

| Demographics and baseline data |  |  |  |
| --- | --- | --- | --- |
| Characteristic | n/N (%) or mean (SD) |  |  |
|  | Overall | Test product | Placebo |
| N (participants) | 43 | 22 | 21 |
| Age (Years) | 27.1 (7.7) | 27.0 (8.1) | 27.2 (7.5) |
| Sex |  |  |  |
| M | 43 (100.0%) | 22 (100.0%) | 21 (100.0%) |
| Body Mass Index | 23.7 (2.6) | 23.6 (2.5) | 23.8 (2.7) |
| Stimulated Leg |  |  |  |
| Left | 22 (51.2%) | 13 (59.1%) | 9 (42.9%) |
| Right | 21 (48.8%) | 9 (40.9%) | 12 (57.1%) |
| Stimulated Leg |  |  |  |
| Dominant | 20 (46.5%) | 11 (50.0%) | 9 (42.9%) |
| Non-Dominant | 23 (53.5%) | 11 (50.0%) | 12 (57.1%) |
| Data are for the Full analysis set (FAS). SD= standard deviation; %=percent. |  |  |  |

### Tolerability and pharmacokinetics

The NAM/PN supplementation was well tolerated, and no adverse events related to the NAM/PN supplementation were reported. Overall, a total of 15 mild adverse events and no serious adverse event were reported (Table 2). One participant contracted a COVID-19 infection during the trial and missed the last visit on Day 8. Other mild adverse events were related to the muscle injury or the muscle biopsy procedures and all were resolved rapidly. NAM/PN supplementation significantly increased serum levels of NAM and PN confirming compliance and uptake of the supplement (Figure 2A-B). Notably, supplementation also resulted in an increase of the bioactive form of PN, pyridoxal-5’-phosphate (PLP) that showed a bioaccumulation effect following 9-days of intake (Figure 2C, NAM/PN group Day 0 vs. Day 8).

**Figure 2:**
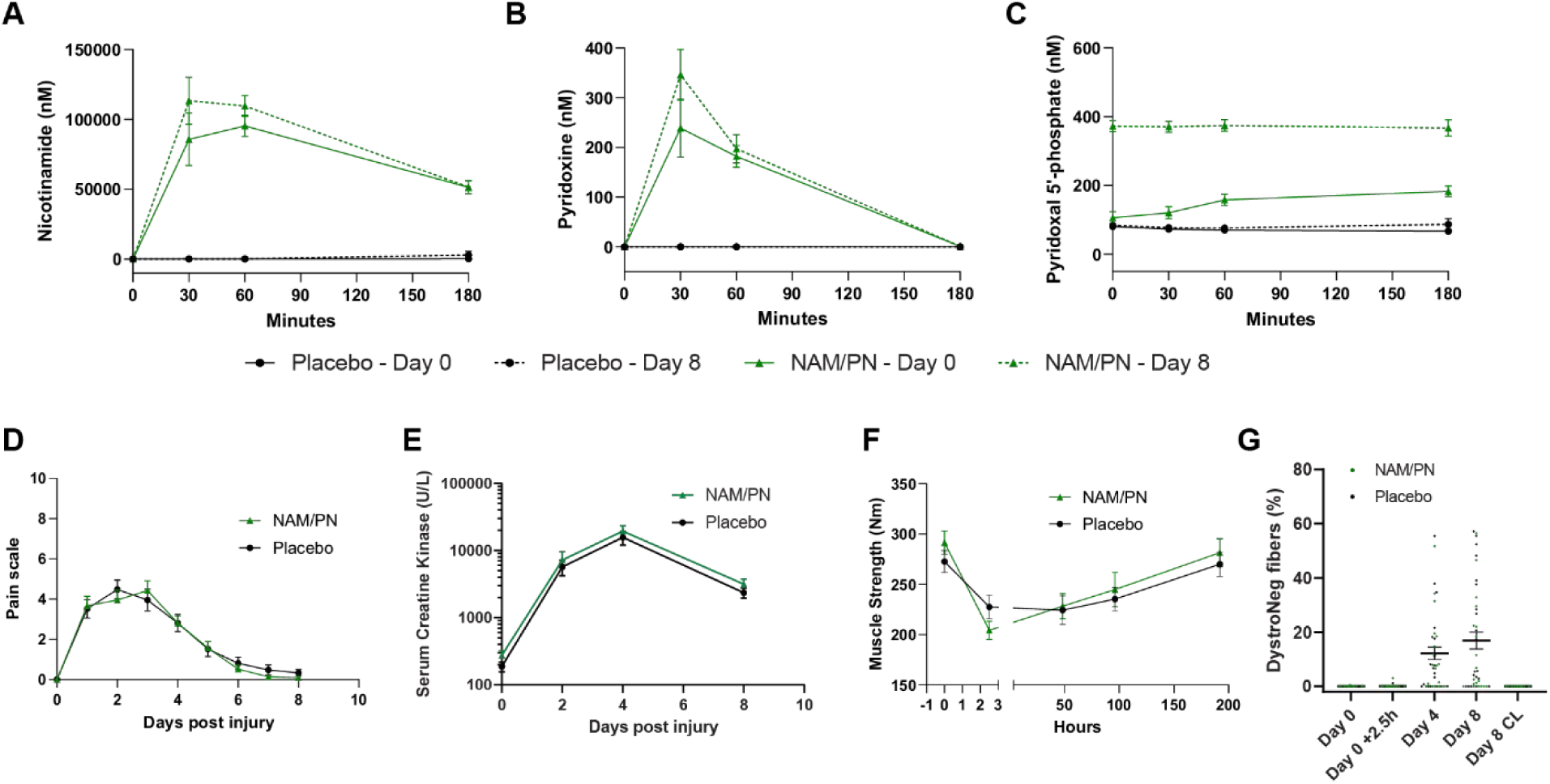
NAM/PN supplementation increases serum levels of NAM and PN and Neuromuscular Electrical stimulation (NMES) protocol elicits a muscle injury. Pharmacokinetic profiles of (A) nicotinamide, (B) pyridoxine and (C) pyridoxal 5’-phosphate (bioactive form of pyridoxine), were assessed on blood samples at Day 0 and Day 8. For each day, blood has been collected at four different timepoints (pre-supplementation and 30 min, 1h, and 3hrs post-supplementation (with NAM/PN or placebo)). (D) Delayed onset muscle soreness (or muscle soreness) was evaluated using a visual analogue scale, which ranges from 0 (normal, no pain) to 10 (extremely painful), at baseline (D0, pre-NMES) and after having induced muscle damage (daily at home by the study participants from Day 1 to Day 8). (E) Serum creatine kinase measurement in blood samples collected for the hematology and serum chemistry assessment at Day 0, Day 2, Day 4 and Day 8. (F) Muscle strength was recorded at baseline (Day 0, pre-NMES) and after having induced muscle damage at Day 0 + 2.5hrs, Day 2, Day 4 and Day 8 in the stimulated leg. Maximal isometric muscle strength was measured in a seated position in a dynamometer at a 70° knee angle (straight leg is zero degrees), as the greatest knee extensor torque measured over 3 attempts. (G) Muscle damage was assessed by the presence of dystrophin-negative (DystroNeg) fibers on cross sections of muscle biopsies collected at baseline (Day 0, contralateral leg) and after having induced a muscle damage at Day 0 (Day 0 + 2.5hrs, stimulated leg), Day 4 (stimulated leg) and Day 8 (stimulated and contralateral legs). (A-C) NAM/PN group, n=21 and 20 at Day 0 and Day 8, respectively; placebo group, n=21 and 20 at Day 0 and Day 8, respectively. (D-F) NAM/PN group, n=21; placebo group, n=21. (G) NAM/PN group, n=19; placebo group, n=20. (A-G) Data are represented as mean ± SEM with, if present, individual dots representing individual subjects.

**Table 2.**

| **Adverse events** (data are on full analysis set) |  |  |  |  |
| --- | --- | --- | --- | --- |
| **Type of adverse event** | **Events in test product group** | **Events in placebo group** | **Severity** | **Relationship to NAM/PN supplementation** |
| **All adverse events** | 9 | 6 |  |  |
| **Serious adverse events** | 0 | 0 |  |  |
| **Infections and infestations** | 1 | 0 |  |  |
| - COVID-19 | 1 | 0 | Mild | Not Related |
| **Injury, poisoning and procedural complications** | 1 | 1 |  |  |
| - Anaesthetic complication | 1 | 1 | Mild | Not Related |
| - Musculoskeletal procedural complication | 1 | 1 | Mild | Not Related |
| **Investigations** | 4 | 1 |  |  |
| - Aspartate aminotransferase increased | 4 | 1 | Mild | Not Related |
| **Musculoskeletal and connective tissue disorders** | 2 | 1 |  |  |
| - Pain in extremity | 2 | 1 | Mild | Not Related |
| **Nervous system disorders** | 1 | 2 |  |  |
| - Headache | 1 | 2 | Mild | Not Related |
| **Renal and urinary disorders** | 0 | 1 |  |  |
| - Haematuria | 0 | 1 | Mild | Not Related |

### NMES protocol induces muscle injury and elicits a regenerative response

Muscle regeneration was induced unilaterally through repeated neuromuscular electrical stimulations (NMES) combined with eccentric contractions of the vastus lateralis muscle. This NMES protocol induced significant clinical signs of muscle damage such as muscle soreness, release of creatine kinase to the circulation, and decrease in quadriceps muscle strength, with no effect of the supplementation on these parameters (Figure 2D-F). Notably, to quantify the extent of muscle damage on cross sections of muscle biopsies, we will rely on the presence of dystrophin-negative fibers (Figure 2G). The proportion of dystrophin-negative at Day 0 + 2.5hrs, Day 4 and Day 8 did not differ significantly between the placebo vs. NAM/PN groups (Figure S1A-C). As demonstrated in previous studies using the same muscle damage model, dystrophin-negative fibers serve as a reliable marker of muscle damage due to their association with the absence of other key structural proteins, significant macrophage infiltration, and the presence of hallmark regenerative indicators (*28, 31–33, 36*). In the present muscle biopsies, we can indeed observe that dystrophin-negative fibers either display massive infiltration of nuclei, sign of ongoing necrosis, or display myogenin^+^ nuclei and expression of eMyHC, hallmarks of active regeneration (Figure S1D, E). In a pilot study, the necrotic state of the dystrophin-negative fibers has been further confirmed using the IgG staining technique which demonstrates that most dystrophin-negative fibers were also IgG positives (Figure S1F).

Only a limited number of studies have explored human muscle regeneration, providing insights into the kinetics of MuSC activation and differentiation. They have demonstrated that MuSCs exhibit active amplification and differentiation approximately 7 to 8 days after muscle injury, with minimal to no activity detected between the first and fourth day (*30, 31, 33, 34*). In line with these previous findings, our analysis of the blinded quantification of MuSC by immunohistofluorescence revealed a similar pattern (Figure 3). At Day 4, no MuSC amplification was observed. A slight unexpected decrease in the number of Pax7^+^ cells was detected compared to baseline, possibly corresponding to the decline of Pax7 after MuSC activation that has been reported in animal models (*37*). A robust MuSC response became evident at Day 8, prompting us to focus subsequent analyses on this time point where the number of Pax7^+^ cells (Figure 3A) and myogenin^+^ cells (Figure 3B) significantly increased compared to the baseline. Furthermore, the appearance of regenerating fibers, indicated by the expression of embryonic myosin (eMyHC, Figure 3C), was also observed at Day 8 and confirmed the regenerative response. These findings emphasize the effective translation of the initial activation of quiescent MuSCs into a genuine regenerative response, ultimately leading to the repair of the damaged muscle fibers.

**Figure 3:**
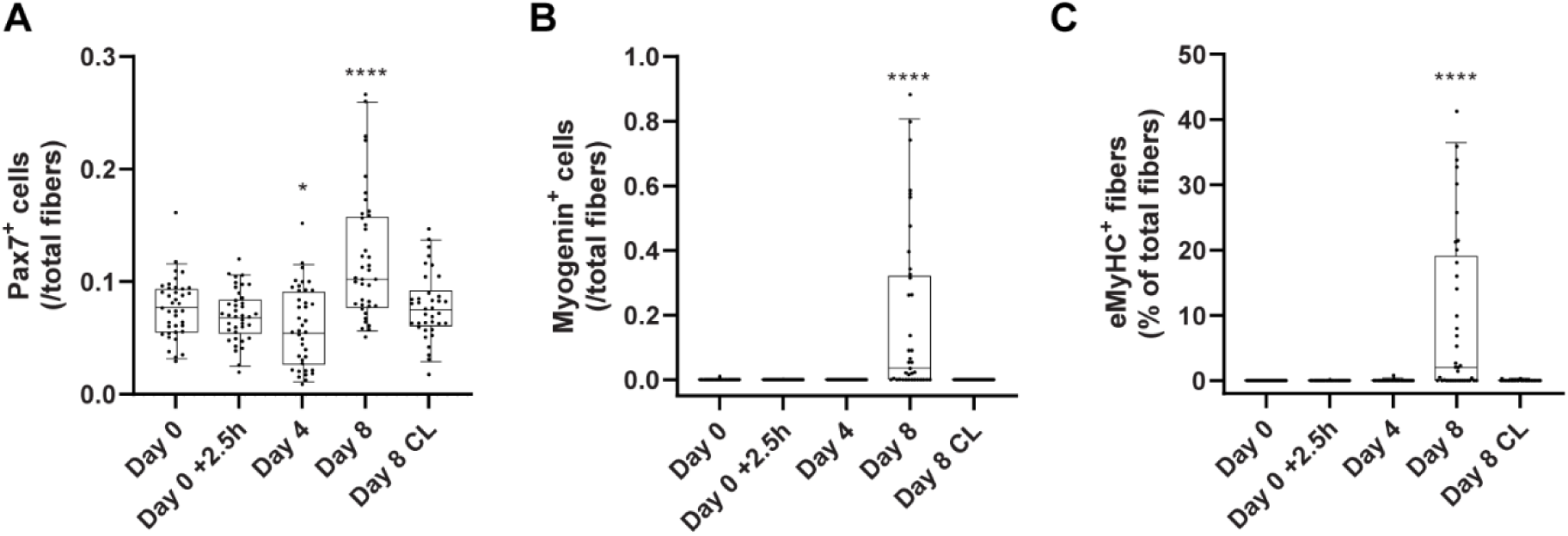
NMES protocol induces a regenerative response. (A-C) Content of Muscle Stem Cell (MuSC) and their progenies were assessed by immunohistofluorescence on cross sections of muscle biopsies collected at Day 0 (pre-NMES, contralateral leg) and at Day 0 + 2.5hrs (stimulated leg), Day 4 (stimulated leg) and Day 8 (stimulated leg and contralateral leg (CL)) after having induced a muscle injury. (A) Activation and proliferation of MuSCs was evaluated by the quantification of Pax7^+^ cells. (B) Differentiation of MuSCs was measured by the number of myogenin^+^ cells and (C) fiber regeneration was assessed by the proportion of fibers displaying immunoreactivity for embryonic myosin (eMyHC). Data are expressed relative to the total number of fibers with individual dots representing individual participants (n=39). *, **** indicated significant difference (p<0.05, p<0.0001, respectively) compared to baseline (Day 0).

### Inter-individual variability in extent of muscle damage affects MuSC outcomes

The extent of muscle damage, as measured by immunohistofluorescence on muscle biopsies by the presence of dystrophin-negative fibers, was highly variable between participants, ranging from 0 to ∼60% of dystrophin-negative fibers (Figure 4A-C,E-G). Notably, about a quarter of the participants showed very low or no sign of muscle damage as observed on the muscle biopsies (from 0 to ∼1% dystrophin-negative fibers, Figure 4C,G, orange dots). These participants did not show a regenerative response as seen by the very low number of MyoD^+^ (Figure 4D) and myogenin^+^ (Figure 4H) cells. The activation of MuSCs and the trigger of a regenerative response were directly linked to the presence of a muscle damage as the percentage of dystrophin-negative fibers strongly correlated with the number of Pax7^+^ cells (Figure 4I, r=0.61), MyoD^+^ cells (Figure 4J, r=0.94), and myogenin^+^ cells (Figure 4K, r=0.97). Based on the direct relationship between these variables and the significant variability observed during the blind data review, we decided to correct for the extent of muscle damage by adjusting the outcomes of interest for the extent of muscle damage via normalization of histology quantifications to the number of dystrophin-negative fibers.

**Figure 4:**
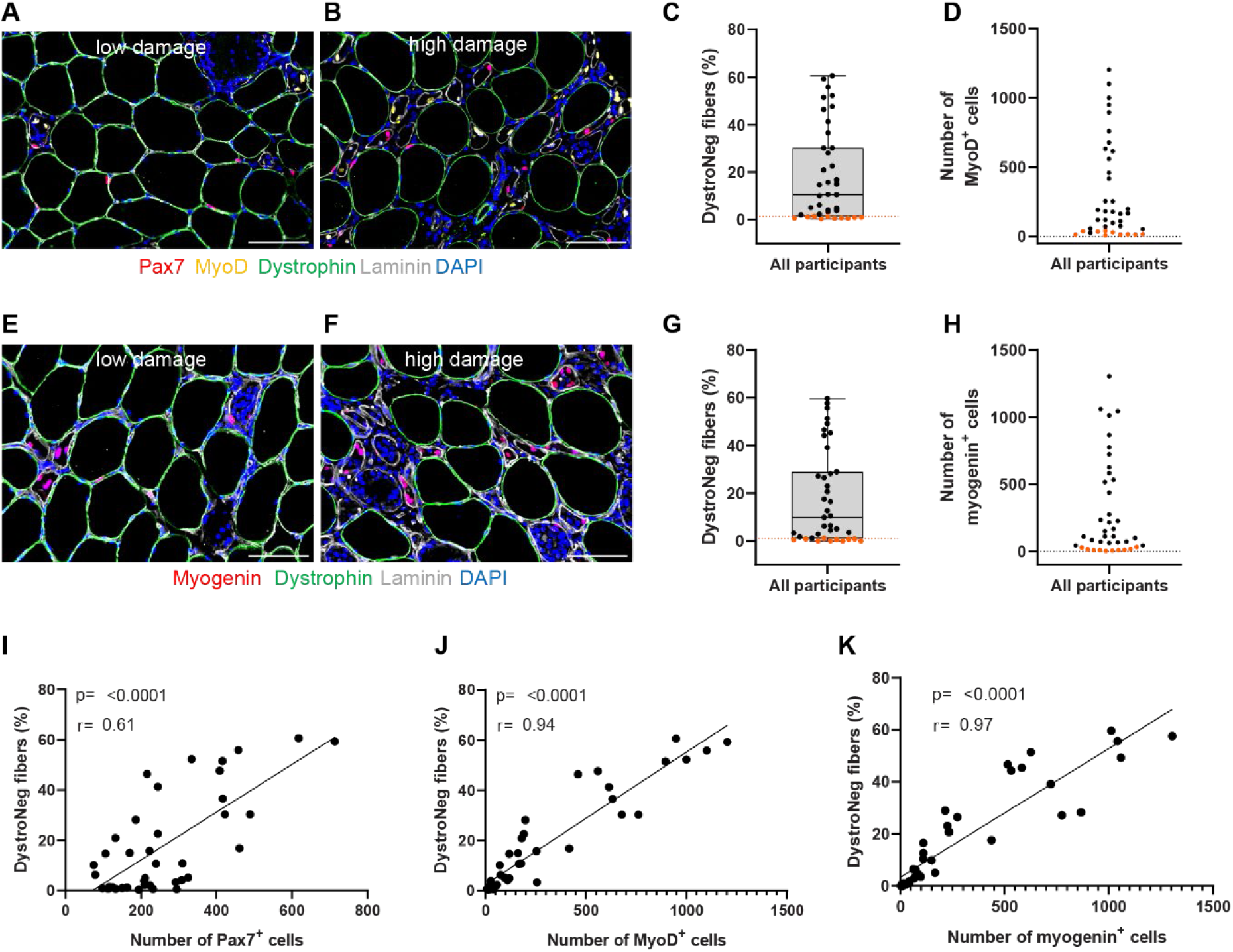
Variability in the extent of muscle damage between subjects is an important covariate that affects MuSC outcomes. Muscle damage as measured by the quantification of dystrophin-negative (DystroNeg) fibers was assessed on the same cross sections used to quantify (A-D) Pax7^+^ and MyoD^+^ cells and (E-H) Myogenin^+^ cells on muscle biopsies collected at Day 8 (stimulated leg). Representative images of a muscle biopsy with low and high level of muscle damage are shown in (A,E) and (B,F), respectively. (C,D,G,H) Orange dots identify subjects at or below the first percentile based on the percentage of dystrophin-negative fibers (with very or no sign of muscle damage). Correlation analyses between the percentage of dystrophin-negative fibers and the number of (I) Pax7^+^ cells, (J) MyoD^+^ cells and (K) myogenin^+^ cells. (C,D,G,H,I-K) Individual dots represent individual participants. Placebo group, n=20; NAM/PN group, n=19. Scale bars = 100µm.

### NAM/PN supplementation increases the amplification and differentiation of MuSCs

To dynamically monitor the different states of myogenesis, we quantified the number of Pax7^+^, MyoD^+^ and myogenin^+^ cells, capturing the main transition from activation, proliferation up to terminal differentiation of MuSCs. Since Pax7^+^ marks both quiescent MuSCs and MuSCs that respond to muscle injury, Pax7^+^ cells were categorized into activated Pax7^+^ cells adjacent to damaged myofibers and quiescent Pax7^+^ cells adjacent to healthy myofibers (Figure S2A). The correlation between muscle damage and activated Pax7^+^ MuSCs (Figure S2B, r=0.96), but not quiescent Pax7^+^ MuSCs (Figure S2C, r=-0.23) validated this classification. Significantly, MuSCs that responded to the muscle injury protocol exhibited a comprehensive advancement in the myogenic program, starting from activated Pax7^+^ cells, progressing to MyoD^+^ cells, and reaching the terminally differentiated state as myogenin^+^ cells. This is illustrated by the strong correlation observed between the number of activated Pax7^+^, MyoD^+^ and myogenin^+^ cells (Figure S2D-F).

The supplementation of NAM/PN significantly increased the number of activated Pax7^+^ cells (Figure 5A,B; +29%; p=0.03 vs placebo), while the number of quiescent Pax7^+^ cells was not affected by the treatment (Figure 5C,D). Furthermore, immunostaining for MyoD revealed a substantial 67% increase in the number of MyoD^+^ cells following NAM/PN vs placebo supplementation (Figure 5E,F). The supplementation of NAM/PN also significantly enhanced the late differentiation process by elevating the number of myogenin^+^ cells by 34% compared to the placebo group (Figure 5G,H). These conclusions were confirmed when analyzing the full set of participants, including those with minimal or no apparent muscle damage (Figure S3A-D). Exploratory analyses with subsets of subjects showing 5% or more damaged fibers and those above the median value for damaged fibers further support our findings. Specifically, all endpoints remained statistically different between the two groups for both subsets (p < 0.05), except the number of MyoD^+^ and Myogenin^+^ cells in the latter subset that demonstrated a clear trend towards significance, with p values of 0.051 and 0.064, respectively (Figure S3E-L). Overall, our findings demonstrate that NAM/PN supplementation specifically promotes the activation, proliferation, and differentiation of MuSCs by increasing the number of activated Pax7^+^ cells, MyoD^+^ cells, and myogenin^+^ cells. This demonstrates that NAM/PN supplementation facilitates the complete progression of different MuSC progenies through the myogenic program without affecting the quiescent MuSC pool.

**Figure 5:**
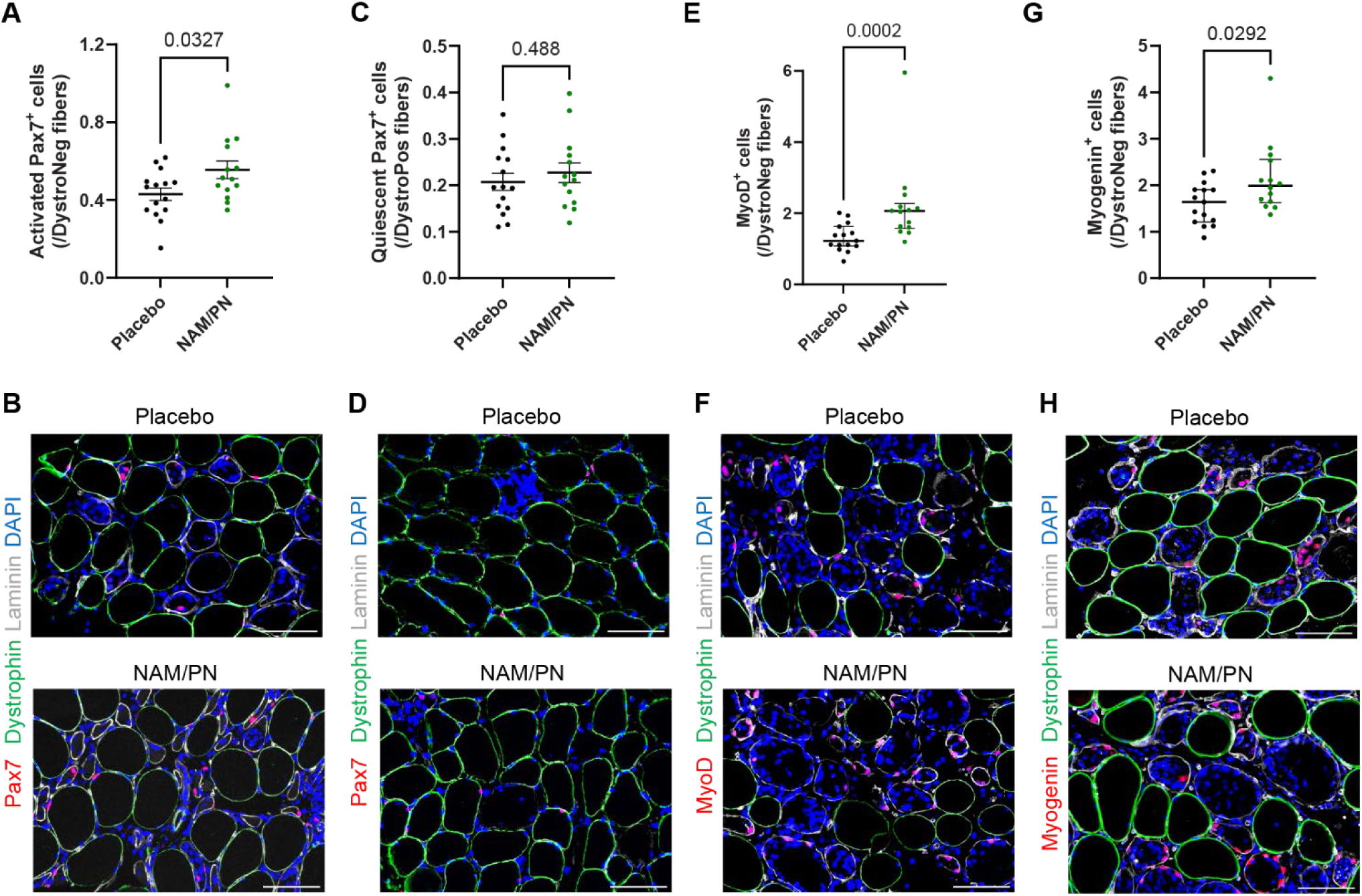
NAM/PN supplementation increases the number of activated Pax7^+^ cells as well as the number of MyoD^+^ and myogenin^+^ cells. The number of MuSCs and their progenies were quantified by immunohistofluorescence on cross section of biopsies collected at Day 8 (stimulated leg) and compared between NAM/PN vs. placebo group. Quantification and representative images of (A,B) activated Pax7^+^ cells; (C,D) quiescent Pax7^+^ cells; (E,F) MyoD^+^ cells, and (G,H) myogenin^+^ cells. Data are represented as (A,C) mean ± SEM or (E,G) median with interquartile range; with individual dots representing individual subjects and are expressed relative to the number of (C) dystrophin-positive (DystroPos) and to the number of (A,E,G) dystrophin-negative (DystroNeg) fibers. All subjects with apparent muscle damage (above the first quartile based on the percentage of dystrophin-negative fibers) on muscle biopsy at Day 8 (stimulated leg) were included in the analyses. Placebo group, n=15; NAM/PN group, n=14. Scale bars = 100µm.

### NAM/PN supplementation accelerates muscle repair

The impact of NAM/PN supplementation on muscle fiber regeneration was assessed by quantifying the expression of embryonic myosin (eMyHC), a well-accepted marker of regenerating fibers (*38*). In order to assess the maturity of regenerating myofibers, damaged fibers were segmented based on the coverage of eMyHC staining, ranging from 0% to 100% of the myofiber area (Figure 6A). Analysis of the distribution profile revealed that approximately 50% of the damaged fibers exhibited no eMyHC expression (Figure 6B), which is consistent with the relatively early time point (Day 8 after injury) and previously published findings (*29*). Furthermore, we observed that the proportion of damaged fibers lacking eMyHC expression was reduced in the NAM/PN group compared to the placebo, while the proportion of fibers beginning to express eMyHC was increased (Figure 6B). Overall, the percentage of damaged fibers expressing eMyHC^+^ at Day 8 was significantly higher by 37% in the NAM/PN group compared to the placebo (Figure 6C,D), indicating that NAM/PN supplementation accelerates the differentiation of MuSCs into eMyHC^+^ regenerating fibers. Similarly to the MuSC readouts (Figure 4I-K), the percentage of eMyHC^+^ fibers exhibited a strong correlation with the extent of muscle damage (Figure S4A). Interestingly, the number of eMyHC^+^ fibers strongly correlated with Pax7^+^, MyoD^+^, and myogenin^+^ cells (Figure S4B-D), confirming that increased MuSC upon NAM/PN treatment directly links with enhanced regeneration. Collectively, our data demonstrate that NAM/PN supplementation accelerates the muscle repair process, as evidenced by the increased proportion of damaged fibers undergoing active regeneration.

**Figure 6:**
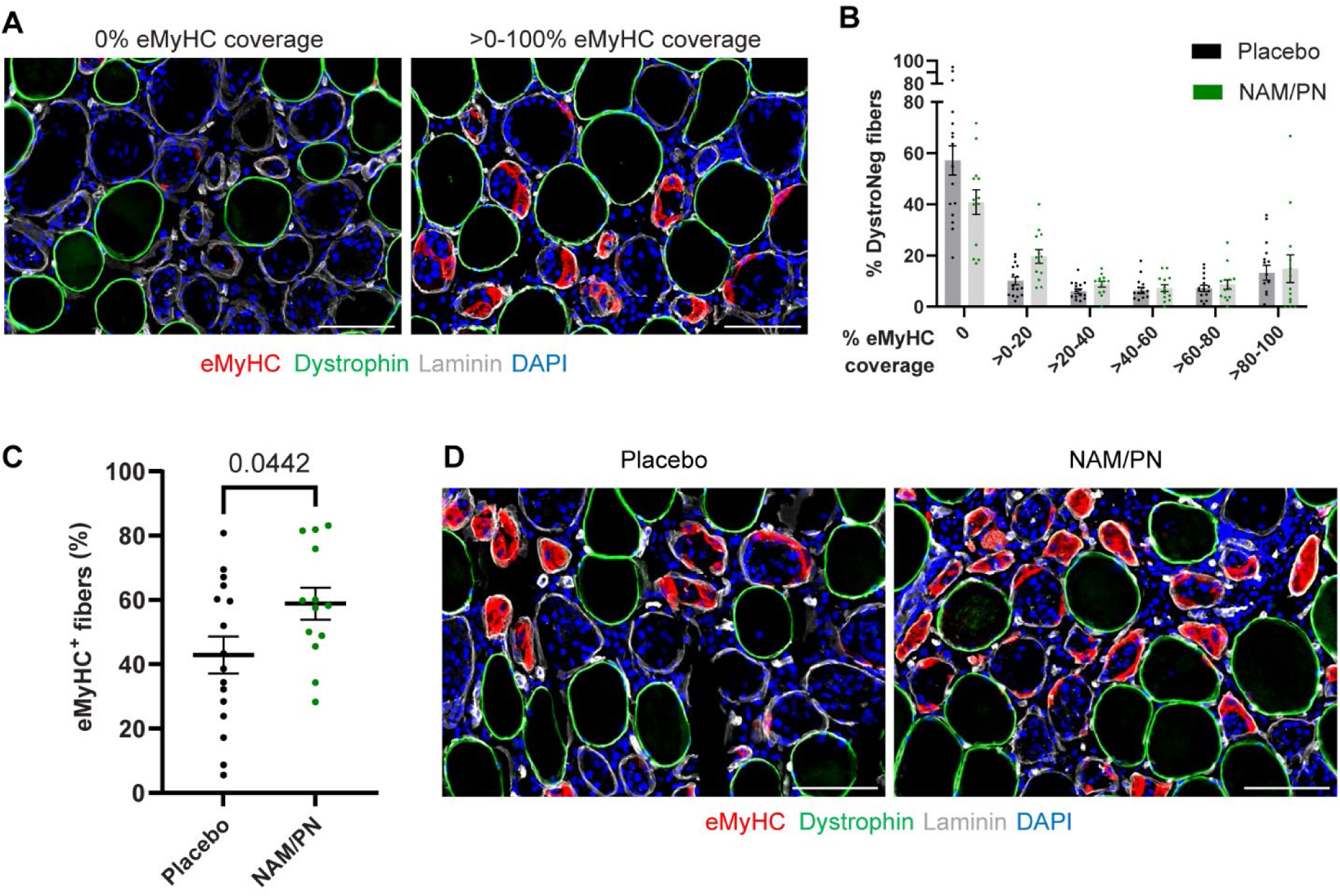
NAM/PN supplementation accelerates the muscle repair process. Fiber regeneration has been evaluated by immunohistofluorescence on cross section of biopsies collected at Day 8 (stimulated leg). (A) Representative images for eMyHC staining depicting examples of damaged fibers with different extent of eMyHC staining ranging from 0% to 100% coverage. (B) Quantification of the area covered by eMyHC staining within each damaged fiber and distribution of damaged fibers based on the percentage of eMyHC coverage. Data are represented as mean ± SEM with individual dots representing individual subjects. (C) The numbers of damaged fibers displaying immunoreactivity for embryonic myosin (eMyHC) were quantified and compared between NAM/PN vs. placebo group. Data are represented as mean ± SEM with individual dots representing individual subjects and are expressed relative to the number of dystrophin-negative fibers. (D) Representative images for eMyHC staining illustrating difference between NAM/PN and placebo groups. (A-D) All participants with apparent muscle damage (above the first quartile based on the percentage of dystrophin-negative fibers) on muscle biopsy at Day 8 (stimulated leg) were included in the analyses. Placebo group, n=16; NAM/PN group, n=13. Scale bars = 100µm.

## DISCUSSION

In this study, we conducted a double-blinded randomized placebo-controlled clinical trial to investigate the efficacy of NAM/PN supplementation on muscle repair after an eccentric contraction-induced muscle damage in healthy men. We consistently observed improvements in the different phases of myogenesis and muscle regeneration, including the amplification of Pax7^+^ activated MuSCs, their transition to MyoD expressing progenitors, and their terminal differentiation to myogenin. Additionally, we observed a positive effect on the maturation of these myogenic progenitors into regenerating myofibers expressing embryonic myosin.

Our findings additionally provide valuable insights into the tolerability and pharmacokinetics of NAM/PN supplementation. The doses of NAM and PN used in our study are within the safety limits defined by the Council for Responsible Nutrition (*39*). Daily oral supplementation with NAM/PN for 9 days did not result in any adverse effects and led to a rapid increase in serum levels of NAM and PN, with Cmax values reached between 30 to 60 minutes. Interestingly, the bioactive form of PN, pyridoxal-5’-phosphate (PLP), that exhibited bioaccumulation between Day 0 and Day 8 reached a plateau after the 9-day intake period. This observation aligns with previous pharmacokinetics studies, which demonstrated that, while a linear relationship between vitamin B6 intake and blood levels of PLP is observed for doses up to 8mg/day (*40, 41*), higher intakes did not show a dose-response progression, and PLP levels reached an upper level steady-state threshold (*42*).

The use of the NMES model with forced lengthening contractions has been confirmed to trigger myofiber necrosis and elicit a robust response from MuSCs, leading to the appearance of regenerating fibers. However, the degree of myofiber injury damage varied significantly among participants. This is likely influenced by individual tolerability to the electric current during stimulation and individual properties of the subcutaneous tissues. Furthermore, human studies rely on small muscle biopsies, which may not capture the full extent of the injury. This important variability poses a challenge in normalizing MuSCs and their progeny. Considering the strong correlation between muscle damage and regeneration, it was crucial to account for the degree of damage to better estimate the potential treatment effects.

NAM, as other forms of vitamin B3, can be converted to NAD^+^, an essential coenzyme for redox reactions central to energy metabolism, and a cofactor or substrate of hundreds of enzymes playing a role in multiple cellular processes (*43*). In our previous preclinical research (*26*), we have discovered that the effect of NAM on MuSCs was independent of NAD^+^ and did not require its conversion to NAD^+^, and signalled through a partial CK1-mediated β-catenin activation. Several NAD^+^ precursors can directly or indirectly generate NAM in vivo after oral intake (*44*). In recent years, a few clinical trials have investigated the effects of different NAD^+^ precursors on skeletal muscle homeostasis and regeneration. One notable study by Jensen et al. explored the impact of nicotinamide riboside (NR) supplementation, in combination with pterostilbene, on muscle regeneration in elderly individuals (*45*). Their findings, primarily evaluating MuSC content measured by flow cytometry, did not demonstrate any significant effect on muscle regeneration. It is important to note that the study by Jensen et al. lacked a direct marker for assessing the extent of muscle damage, which limited their ability to evaluate the isolated effect on damaged fibers. Furthermore, the lower blood creatine kinase levels observed two days after the contraction protocol in their study (approximately an average of 1000 U/L) compared to our study (approximately 6000 U/L) suggest that the extent of myofiber damage was less substantial. Another study investigated the long-term effects of NR supplementation on mitochondrial biogenesis in muscle and fat tissue (*46*). They reported a beneficial effect on MuSC differentiation showing a change in the ratio of Pax7 expression over the late myogenic differentiation marker myogenin. However, this study did not involve muscle injury and MuSCs readouts were exploratory and mostly deduced from gene expression analyses on whole muscles or cultured myoblasts. Overall, direct comparisons with the studies by Jensen et al. and Lapatto et al. are precluded due to differences in supplementation approaches and variations in evaluation methods. Moreover, we have shown in a mouse model that the levels of NAM resulting from NR conversion were 50 times lower compared to the levels achieved through direct NAM supplementation, failing to reach the threshold at which NAM exhibits its active effects on MuSCs (*26*). Additionally, the MuSC and muscle repair efficacy in the present study not only relies on NAM but on the combination of NAM with PN (*26*).

Currently, there are no treatments specifically targeting MuSCs for promoting muscle repair and recovery that are implemented in clinical practice. Most of the available experimental solutions are at the preclinical stage, with only a limited number of clinical studies conducted. Notably, testosterone has been shown to stimulate MuSC proliferation in humans (*47*) (*48*), but it is not without side effects and safety considerations are crucial when developing treatment approaches for promoting muscle regeneration across different age groups and severity of cases. Nutritional solutions may offer a safer solution. The role of dietary protein in supporting muscle adaptation and growth following exercise in adults has been widely demonstrated (*49, 50*). This beneficial effect has been primarily attributed to the well-established impact on myofibrillar protein synthesis (*51, 52*) with few studies looking at a potential influence on MuSC response (*53*)(*54*). Therefore, our findings on the positive effect of NAM/PN supplementation on MuSCs and fiber regeneration have direct opportunities for clinical translation for the management of sport-related muscle damage, where boosting regeneration can accelerate the repair process and could be combined with dietary protein strategies for enhanced muscle recovery.

Our findings could also have implications for a wide range of muscle disorders in which a compromised MuSC function and muscle regeneration is a pathological hallmark. Notably, sarcopenia, the age-related loss of muscle mass, strength, and function, is in part characterized by a decline in MuSC function and number, resulting in compromised regenerative capacity of the skeletal muscle (*14, 55–57*). A clinical study has shown that older individuals struggle to recover from disuse-induced atrophy and fail to expand the MuSC pool during rehabilitation (*58*). Our previous preclinical investigations including rodent models and primary myoblasts from aged humans have demonstrated that NAM/PN overcomes MuSC dysfunction and regenerative failure associated with aging (*26*). Taken together, these findings strongly support the potential of NAM/PN as a novel nutritional intervention for alleviating age-related muscle decline. Another application is cancer cachexia which results from systemic inflammation and an imbalance between protein synthesis and degradation (*59*), coupled to myofiber damage and impaired regeneration in the muscle wasting process (*16, 17, 60*). Interestingly, using mouse models of cancer cachexia, two studies have shown that supplementation with niacin could mitigate muscle wasting (*61, 62*) but the potential role of MuSCs was not investigated. Muscular dystrophies and myopathies encompass a wide and heterogeneous group of genetic conditions characterized by muscle weakness (*63*). Altered MuSC function and muscle regeneration capacity is a common hallmark of several muscular dystrophies and myopathies (*64*). Notably, targeting MuSCs in the context of Duchenne Muscular Dystrophy (DMD) holds great potential in mitigating the dystrophic phenotype (*65–67*). Several pharmacological drugs that affect MuSC function have demonstrated efficacy in preclinical models of DMD (*68–70*). Additionally, while MuSCs play a central role in muscle regeneration, muscle maintenance and repair require a highly orchestrated coordination between multiple cell types and their microenvironment (*71*). For example, MuSCs have been shown to secrete exosomes that regulate collagen biosynthesis in fibrogenic cells to prevent excessive extracellular matrix deposition (*72*). Fibrosis being a hallmark of many muscle diseases, these results highlight a potential benefit of promoting MuSC function beyond the formation of healthy myofibers.

Although our study did not directly investigate these specific disorders, the observed positive effects of NAM/PN supplementation in healthy volunteers may have potential benefits for individuals affected by these conditions. Further research is needed to explore the specific mechanisms and implications of NAM/PN supplementation in these muscle disorders.

## MATERIALS AND METHODS

### Study Design and Participants

This randomized, double-blind, placebo-controlled study was approved by the Regional Scientific Ethical Committees of Copenhagen (protocol number H-20081080). All participants provided written consent to participate, after the nature and possible consequences of the studies were explained, and all procedures conformed to the Declaration of Helsinki. This single-center, randomized, double-blind, placebo-controlled trial was conducted in the Institute of Sports Medicine Copenhagen between April 2021 and May 2022. This study is registered on ClinicalTrials.gov with the identifier NCT04874662. The sample size calculation was based on a variability of 20% (*31*) and an effect size of 30%. To reach a statistical power of 80%, if all the assumptions made were met, 20 subjects were needed per group (40 subjects in total).

Before inclusion all participants were screened by a physician and deemed overall healthy based on blood samples, urine analysis, blood pressure measurement, EKG, neurological exam, medical history and an interview. All participants were men, aged between 18-50 with a BMI between 18.5 and 24.9 kg/m^2^ and had normal dietary habits. Exclusion criteria included current smoking, participation in structured strength exercise less than 3 months prior to participation in the study, use of corticosteroids, anabolic steroids, growth hormones, anti-coagulant or anti-aggreging agents or having participated in a clinical study in the last 3 months. Consumption of supplements containing vitamin B3 and/or vitamin B6 was also prohibited at least two weeks before the inclusion and during the intervention, other than those provided by the investigator. Eligible participants were randomized in a 1:1 ratio to supplementation (NAM/PN) or control group (placebo), on Day 0 by an investigator not involved in the inclusion process. The randomization was stratified by age (18≤age<35 and 35≤age≤50). Inclusion of participants from both groups was similarly distributed throughout the study period.

### NAM/PN and placebo administration

Participants received, in a double-blinded manner, oral supplementation with NAM/PN (714 mg NAM + 19 mg PN, DSM Nutritional Products GmbH, Germany) or placebo (microcrystalline cellulose, Hänseler AG, Herisau, Switzerland) as two capsules per day to be taken in the morning in fasting (overnight sleep) condition for 9 days (from Day 0 to Day 8).

### Research objectives and prospective selection of endpoints

The primary objective of this study was to investigate if a combination of nicotinamide (NAM) and pyridoxine (PN) intake would improve human skeletal muscle regeneration, through enhanced activation of MuSCs. As stated in the trial registration (NCT04874662), the primary endpoint was prospectively selected as the number of MuSCs (Pax7 and myogenin) on sections of muscle biopsies from the stimulated leg, comparing the study intervention (NAM/PN) to the control intervention (placebo).

### Muscle injury protocol

One leg, dominant or non-dominant, was randomly selected for neuromuscular electrical muscle stimulation (NMES) coupled with eccentric contractions to induce muscle damage. The electrical stimulation was applied to the vastus lateralis with a CefarCompex mi-Theta 600 Muscle stimulator (DJO Nordic, Malmoe, Sweden). Adhesive electrodes were placed on shaved and cleaned skin. A 10 x 5 cm adhesive electrode was placed proximal (inguinal area) on the vastus lateralis. And two 5x5 cm adhesive electrodes were placed distally on the vastus lateralis, spaced out so the entire width of the vastus lateralis was covered. After placements of the adhesive electrodes the participants did a 5 min warm up of slow to moderate cycling on a bike ergometer, followed by 5-10 min familiarization with the electric stimulation, where the electric current was gradually increased. The stimulation was delivered every 15 seconds with a frequency of 35 Hz, 300 μs pulse-duration, 0-120 mA, and a 0,75 second ramp-up/down period.

The stimulations were performed in a Kin-Com dynamometer (Kin-COM, model 500-11, Kinetic Communicator, Chattecx, Chattanooga, TN), in which each stimulation was accompanied by mechanical flexion of the knee. A total of 5 x 20 slow (3.5 second stimulations and flexion with an angular velocity of 30° pr. second) and 5 x 20 fast (1.5 seconds stimulation and flexion with an angular velocity of 180° pr. second) stimulations. The stimulation current was continuously increased till the participants tolerations threshold. This method was slightly adapted from previous used protocols (*31, 32*).

### Blood samples

Blood samples were collected at Day 0, Day 2, Day 4 and Day 8 from an antecubital vein into EDTA tubes and placed on ice until storage at –80°C. Blood analysis of creatine kinase was carried out at the Clinical Biochemistry department at Bispebjerg and Frederiksberg Hospital.

### Pharmacokinetic assessments

NAM, PN and related metabolites pharmacokinetic profiles were assessed on blood samples at Day 0 and Day 8. For each day, blood will be collected at four different timepoints (pre-supplementation, and 30 min, 1h, and 3hrs post-supplementation). Participants were in fasting conditions until the 3hrs post-supplementation timepoint for blood collection. Serum was rapidly processed and stored at -80°C. Concentrations of nicotinamide (NAM), pyridoxal 5’-phosphate (PLP, bioactive from of PN) and pyridoxine (PN) were measured using LC-MS/MS by BEVITAL (Bergen, Norway; www.bevital.no). Measurements were performed by mixing samples with labelled internal standards, separation on a C8 liquid chromatography column by a gradient-type mobile phase and detecting analytes using electrospray ionization tandem mass spectrometry as described in (*73*)

### Muscle biopsies

Skeletal muscle biopsies were obtained from the vastus lateralis muscles using the percutaneous needle biopsy technique of Bergström (*74*). 1% lidocaine (amgros I/S, Copenhagen, Denmark) was applied subcutaneously as local anaesthetic, and tissue was extracted with a 5 mm diameter biopsy needle and manual suction. The biopsies were spaced by a minimum of 2 cm. Each biopsy was divided with one main portion being embedded in Tissue-Tek (Sakura Finetek Europe, Zoeterwoude, the Netherlands) and frozen in isopentane, precooled by liquid nitrogen for cryosectioning and immunohistofluorescence analyses and the remaining smaller portion being snap frozen in liquid nitrogen. All muscle tissue was stored at –80 °C.

### Muscle soreness

Muscle soreness will be evaluated using a visual analogue scale (VAS), which ranges from 0 (normal, no pain) to 10 (extremely painful). Participants were instructed to slowly sit down unassisted (5 seconds) onto a chair, at home, first thing in the morning, and note on a scale of 1-10 how painful the front thigh muscles (of the stimulated leg) were during this movement. Soreness was daily assessed starting the morning before muscle injury until Day 8.

### Muscle strength test

Maximal voluntary isometric strength (MVIC) was tested by measuring peak isometric torque at a 70° knee flexion in an isokinetic dynamometer (Kinetic Communicator model 500-11). The best of three attempts was reported. The tests were performed immediately before and after the injury protocol, as well as on Day 2, Day 4 and Day 8. This test measures the strength of the whole quadriceps muscle which consists of four individual muscles. It is used as an early marker of muscle damage to assess the effectiveness of the injury protocol, but its sensitivity is limited because only one out of the four muscles involved in the strength test is subjected to electrical stimulation and thus damaged (vastus lateralis).

### Immunohistofluorescence

Frozen tissues were sectioned at 10µm with a cryostat (Leica Biosystems) at −20 °, and sections were placed on glass slides (Superfrost Plus), which were stored at −80 °C before being stained.

All biopsy timepoints were stained for Pax7 (DSHB, Pax7, supernatant, 1:100), myogenin (DSHB, F5D, supernatant, 1:100), eMyHC (DSHB, F1.652, supernatant, 1:100) and dystrophin (Sigma, D8168, 1:100), each one in combination with laminin (Sigma, L9393, 1:200). Primary antibodies were applied overnight to fixed (myogenin; Histofix, 10 min), or unfixed (dystrophin, eMyHC), sections according to the respective antibody datasheets. Sections were then incubated in a cocktail of two secondary antibodies (Thermo Fischer Scientific A-11029, A-11036) for 45 min before mounting in ProLong Gold Antifade Reagent, containing DAPI (Invitrogen, P36931). Dystrophin and eMyHC sections were fixed (Histofix, 10 min) before mounting. For Pax7, sections were fixed in 4% PFA (5 mins) before overnight incubation with antibodies against Pax7, laminin and myosin heavy chain I (DSHB, BA-D5, 1:00), visualized by three secondary antibodies (Thermo Fischer Scientific A-21121, A-21144, A-21076). The DSHB Hybridoma products F1.652, F5D, Pax7, and BA-D5 were deposited to the DSHB by H.M. Blau, W.E. Wright, A. Kawakami, S. Schiaffino, respectively. These sections were imaged with the AxioScan 7 or Z1 slide scanner and images were analysed by manually counting positive (or, for dystrophin, negative) cells in QuPath.

For a more detailed analysis of the Day 8 samples, Pax7/MyoD/dystrophin co-stainings and myogenin/dystrophin co-stainings, slides were stained with various combinations of antibodies against laminin (Sigma, L9393, 1:1000) , dystrophin (Sigma, D8168, 1:500) and Pax7 (DSHB, purified, 2.5 µg/ml, 1:250); MyoD (Abcam, ab133627, 1:500); myogenin (Abcam, ab124800, 1:500); and Hoechst 33342 (Sigma, B2261). All antibodies are diluted in blocking buffer containing 4% BSA (Jackson ImmunoResearch, 001-000-162) in PBS. Slides were fixed with 4% PFA (EMS, #157-4-100), for 15 minutes and permeabilized in cold methanol (VWR, #1.06035.2500) for 6 minutes. Antigen retrieval was performed with two successive incubations of hot 0.01 M pH 6 citric acid during 4 min, and sections were further blocked in 4% BSA for 2 h. To avoid technical issues like cross reaction, interaction, nuclear markers were stained and detected first. Slides were sequentially incubated overnight with primary antibodies anti-Pax7, anti-MyoD, or anti-myogenin at 4°C. Before new incubation, slides were washed three times in PBS/0.1% Triton X-100. Pax7 signal was further amplified using a goat-anti mouse IgG1-biotin (Jackson ImmunoResearch, 115-065-205, 1:1000) followed by conjugation with Streptavidin Alexa555 (Life Technologies, #S-21381, 1:2000), and other antibodies were detected with their specific secondary antibodies (Thermo Fisher Scientific, A32733), while nuclei were detected with Hoechst 33342 (1:10000). Then, the slides were stained for fiber structures. Slides were incubated with primary antibodies anti-laminin, anti-dystrophin for 3 hours at room temperature. Finally, primary antibodies were detected using anti-rabbit IgG (Life Technologies, A-21039, 1:2000) and anti-mouse IgG2b (Life Technologies, A-21141, 1:2000).

For eMyHC/dystrophin co-staining and IgG staining, slides were stained with various combinations of antibodies against laminin (Sigma, L9393, 1:1000), dystrophin (Sigma, D8168, 1:500), human IgG (Beckman Coulter, A79391) and eMyHC (DSHB, F1.652, 1:500); and Hoechst 33342 (Sigma, B2261). All antibodies are diluted in blocking buffer containing 4% BSA (Jackson ImmunoResearch, 001-000-162) in PBS. After 15 min of rehydration in PBS, slides were blocked for 45 min at room temperature in 4% BSA. Slides were stained for 3 h at room temperature using anti-laminin and anti-eMyHC antibodies. Sections were then incubated for 1 h with secondary antibodies (Thermo Fisher Scientific, A-21127, A-21039, A-21127, 1:2000) at room temperature and counterstained with Hoechst (1:10000). Then, dystrophin staining was performed as described previously.

All slides were then mounted using Dako fluorescent mounting medium (Agilent, #S302380-2).

### Imaging and quantification

All biopsy sections were entirely scanned with an Olympus VS200 slide scanner at 20× magnification and stitched to 1 image. Quantifications of Pax7^+^, MyoD^+^, myogenin^+^ cells were determined by manually counting on entire muscle sections with researchers blinded to the experimental groups. Quiescent Pax7^+^ cells were distinguished as Pax7^+^ cells belonging to dystrophin-positive fibers and residing under the basal lamina whereas activated Pax7^+^ cells were distinguished as being located inside or in close proximity to dystrophin-negative fibers.

Fibers were detected across the entire sections by an automated image processing algorithm developed internally using the QuPath software and the Fiji open-CSAM tool. Fibers are detected and classified based on both dystrophin and laminin staining. A quality control was performed to remove artifacts. Missing fibers were manually drawn and assigned to the correct class. The number of uninjured fibers was determined from the sections as being dystrophin- and laminin-positive, whereas damaged fibers were detected as being laminin-positive and dystrophin-negative. To quantify the extent of myofiber regeneration, sections were stained for embryonic myosin heavy chain (eMyHC). After cross sectional analysis, the total number of regenerating fiber (eMyHC^+^) and the area covered by eMyHC into the damaged fibers were quantified by applying a threshold analysis to the whole image via Qupath software (pixel classification option).

### Statistics

All data were analyzed and plotted using R version 4.2.2 and GraphPad Prism 9.0 software. Normality of data was determined using Shapiro Wilk normality test. Comparisons were performed using Welch’s t-test for normally distributed data or Wilcoxon rank sum test (equivalent to the Mann-Whitney test) for non–normally distributed data, corresponding paired tests were performed whenever appropriate. Normally distributed data are represented as mean ± SEM and non-normally distributed data as median with interquartile range. Multiple comparisons were performed using a Kruskal-Wallis test with Dunn’s post hoc test or a Friedman test followed by pairwise Wilcoxon signed rank tests to identify different groups and Bonferroni corrections for multiple testing. Spearman’s correlation analyses were conducted for associations between continuous variables. The per-protocol analysis comprised all 39 participants who completed the study.

Because activated MuSCs and muscle regeneration cannot be quantified in the absence of muscle damage, a subgroup statistical analysis was conducted with exclusion of participants with very low muscle damage. Very low muscle damage was defined as the first quartile of the dystrophin-negative fiber distribution, corresponding to 0.74-1.38% of dystrophin-negative fibers (for each readout the sample size is indicated in the respective figure legend). Raw data for the number of fibers, dystrophin-negative fibers, quiescent Pax7^+^ cells, activated Pax7^+^ cells, MyoD^+^ cells and eMyHC^+^ fibers are presented in Table 3. All statistical analyses were 2 tailed, and P values of 0.05 or less were considered statistically significant.

**Table 3:** Raw data for the number of fibers, dystrophin-negative fibers, quiescent Pax7^+^ cells, activated Pax7^+^ cells, MyoD^+^ cells and eMyHC^+^ fibers.

|  |  |  | Placebo group |  |  |  |  | NAM/PN group |  |  |  |  |
| --- | --- | --- | --- | --- | --- | --- | --- | --- | --- | --- | --- | --- |
|  |  |  | Average | SD | Median | Min | Max | Average | SD | Median | Min | Max |
| Pax7/MyoD analyses | Per-protocol population | Total fibers | 1225 | 458 | 1090 | 509 | 2099 | 987 | 477 | 913 | 324 | 2063 |
|  |  | Dystrophin-negative fibers | 243 | 255 | 150 | 5 | 830 | 151 | 204 | 43 | 5 | 639 |
|  | Subjects with apparent muscle damage (>1<sup>st</sup> quartile based on % dystrophin-negative fibers) | Total fibers | 1165 | 354 | 1086 | 753 | 2014 | 1004 | 517 | 973 | 324 | 2063 |
|  |  | Dystrophin-negative fibers | 321 | 249 | 196 | 40 | 830 | 203 | 216 | 82 | 21 | 639 |
| Myogenin analyses | Per-protocol population | Total fibers | 1134 | 469 | 977 | 500 | 2034 | 926 | 409 | 885 | 280 | 1819 |
|  |  | Dystrophin-negative fibers | 218 | 224 | 119 | 0 | 687 | 130 | 174 | 43 | 0 | 514 |
|  | Subjects with apparent muscle damage (>1<sup>st</sup> quartile based on % dystrophin-negative fibers) | Total fibers | 1039 | 327 | 951 | 630 | 1874 | 931 | 448 | 905 | 280 | 1819 |
|  |  | Dystrophin-negative fibers | 288 | 217 | 246 | 29 | 687 | 175 | 183 | 71 | 10 | 514 |
| eMyHC analyses | Per-protocol population | Total fibers | 1188 | 428 | 988 | 660 | 1925 | 973 | 452 | 975 | 290 | 1884 |
|  |  | Dystrophin-negative fibers | 233 | 247 | 124 | 4 | 755 | 137 | 185 | 46 | 1 | 560 |
|  | Subjects with apparent muscle damage (>1<sup>st</sup> quartile based on % dystrophin-negative fibers) | Total fibers | 1107 | 380 | 971 | 660 | 1925 | 964 | 499 | 975 | 290 | 1884 |
|  |  | Dystrophin-negative fibers | 290 | 245 | 193 | 23 | 755 | 197 | 198 | 73 | 11 | 560 |

| Subjects with apparent muscle damage (>1st quartile based on % dystrophin-negative fibers) |  |  |  |  |  |  |  |  |  |  |  |  |
| --- | --- | --- | --- | --- | --- | --- | --- | --- | --- | --- | --- | --- |
|  |  |  | Placebo group |  |  |  |  | NAM/PN group |  |  |  |  |
|  |  |  | Average | SD | Median | Min | Max | Average | SD | Median | Min | Max |
| Pax7/MyoD analyses | quiescent Pax7+ cells |  | 165 | 71 | 159 | 82 | 293 | 165 | 77 | 173 | 61 | 277 |
|  | activated Pax7+ cells |  | 148 | 141 | 96 | 14 | 513 | 124 | 143 | 43 | 10 | 457 |
|  | MyoD+ cells |  | 449 | 394 | 191 | 26 | 1203 | 346 | 327 | 227 | 53 | 949 |
| Myogenin analyses | Myogenin+ cells |  | 474 | 413 | 274 | 67 | 1305 | 322 | 331 | 163 | 43 | 1043 |
| eMyHC analyses | eMyHC+ fibers |  | 163 | 181 | 82 | 2 | 610 | 153 | 169 | 45 | 5 | 425 |

## List of Supplementary Materials

**Figure S1.**

Dystrophin-negative fibers identify damaged and regenerating fibers.

**Figure S2.**

The muscle injury protocol triggers a complete progression of different MuSC progenies through the myogenic program.

**Figure S3**

NAM/PN supplementation increases the number of activated Pax7^+^ cells as well as the number of MyoD^+^ and myogenin^+^ cells.

**Figure S4**

Evaluation of fiber regeneration by immunohistofluorescence on cross sections of biopsies collected at Day 8 (stimulated leg).

## Data File S1

Raw data Excel File.

## Data Availability

All data produced in the present study are available upon reasonable request to the authors

## Acknowledgments

We acknowledge the Core Facility for Integrated Microscopy, Faculty of Health and Medical Sciences, University of Copenhagen, for use of the AxioScan slide scanner.

## Author contributions

Designed and experimental strategy: GH, JM, LGK, MK, JNF, PS, and ALM

Interpretation of results, and writing the manuscript: GH, JM, JNF, PS, and ALM

Performed experiments: GH, JM, AD, KK, SK, BWH, WH

Contributed to experimental strategy and data interpretation: MK, EPM, OEJ, LGK

Analyzed data: EM, GH, JM, JNF, PS, ALM

Conceived and lead the project: JNF, PS

Co-first authors share primary responsibility in conducting experiments, analyses, and interpretation of results for this study. The order of co-first authors reflects the contribution to writing and editing of the manuscript: GH, JM

All authors read and approved the final manuscript.

## References and Notes

1. N. A. Dumont, C. F. Bentzinger, M. C. Sincennes, M. A. Rudnicki, Satellite Cells and Skeletal Muscle Regeneration. Compr Physiol 5, 1027–1059 (2015).

2. J. F. Bachman et al., Prepubertal skeletal muscle growth requires Pax7-expressing satellite cell-derived myonuclear contribution. Development 145, (2018).

3. T. Delhaas, S. F. Van der Meer, G. Schaart, H. Degens, M. R. Drost, Steep increase in myonuclear domain size during infancy. Anatomical record 296, 192–197 (2013).

4. L. B. Verdijk et al., Satellite cells in human skeletal muscle; from birth to old age. Age (Dordr) 36, 545–547 (2014).

5. B. Blaauw, C. Reggiani, The role of satellite cells in muscle hypertrophy. J Muscle Res Cell Motil 35, 3–10 (2014).

6. T. Snijders et al., Satellite cells in human skeletal muscle plasticity. Front Physiol 6, 283 (2015).

7. M. Schmidt, S. C. Schuler, S. S. Huttner, B. von Eyss, J. von Maltzahn, Adult stem cells at work: regenerating skeletal muscle. Cell Mol Life Sci 76, 2559–2570 (2019).

8. T. A. Järvinen, T. L. Järvinen, M. Kääriäinen, H. Kalimo, M. Järvinen, Muscle injuries: biology and treatment. Am J Sports Med 33, 745–764 (2005).

9. T. A. Jarvinen, M. Kaariainen, M. Jarvinen, H. Kalimo, Muscle strain injuries. Curr Opin Rheumatol 12, 155–161 (2000).

10. J. M. Beiner, P. Jokl, Muscle contusion injuries: current treatment options. J Am Acad Orthop Surg 9, 227–237 (2001).

11. P. Edouard et al., Traumatic muscle injury. Nat Rev Dis Primers 9, 56 (2023).

12. T. Laumonier, J. Menetrey, Muscle injuries and strategies for improving their repair. J Exp Orthop 3, 15 (2016).

13. G. Hojfeldt et al., Fusion of myofibre branches is a physiological feature of healthy human skeletal muscle regeneration. Skelet Muscle 13, 13 (2023).

14. H. M. Blau, B. D. Cosgrove, A. T. Ho, The central role of muscle stem cells in regenerative failure with aging. Nat Med 21, 854–862 (2015).

15. V. E. Baracos, L. Martin, M. Korc, D. C. Guttridge, K. C. H. Fearon, Cancer-associated cachexia. Nat Rev Dis Primers 4, 17105 (2018).

16. M. Bossola, E. Marzetti, F. Rosa, F. Pacelli, Skeletal muscle regeneration in cancer cachexia. Clin Exp Pharmacol Physiol 43, 522–527 (2016).

17. W. A. He et al., NF-kappaB-mediated Pax7 dysregulation in the muscle microenvironment promotes cancer cachexia. J Clin Invest 123, 4821–4835 (2013).

18. D. M. D’Souza, D. Al-Sajee, T. J. Hawke, Diabetic myopathy: impact of diabetes mellitus on skeletal muscle progenitor cells. Front Physiol 4, 379 (2013).

19. M. P. Krause et al., Impaired macrophage and satellite cell infiltration occurs in a muscle-specific fashion following injury in diabetic skeletal muscle. PLoS One 8, e70971 (2013).

20. N. Yanay, M. Rabie, Y. Nevo, Impaired Regeneration in Dystrophic Muscle-New Target for Therapy. Front Mol Neurosci 13, 69 (2020).

21. F. Relaix et al., Perspectives on skeletal muscle stem cells. Nat Commun 12, 692 (2021).

22. C. M. Bleakley, P. Glasgow, D. C. MacAuley, PRICE needs updating, should we call the POLICE? Br J Sports Med 46, 220–221 (2012).

23. B. Dubois, J. F. Esculier, Soft-tissue injuries simply need PEACE and LOVE. Br J Sports Med 54, 72–73 (2020).

24. A. L. Mackey, U. R. Mikkelsen, S. P. Magnusson, M. Kjaer, Rehabilitation of muscle after injury - the role of anti-inflammatory drugs. Scand J Med Sci Sports 22, e8–14 (2012).

25. K. M. Morelli, L. B. Brown, G. L. Warren, Effect of NSAIDs on Recovery From Acute Skeletal Muscle Injury: A Systematic Review and Meta-analysis. Am J Sports Med 46, 224–233 (2018).

26. S. Ancel et al., Nicotinamide and Pyridoxine Stimulate Muscle Stem Cell Expansion and Enhance Regenerative Capacity during Aging. Journal of Clinical Investigation, e163648 (2024).

27. D. Hardy et al., Comparative Study of Injury Models for Studying Muscle Regeneration in Mice. PloS one 11, e0147198 (2016).

28. A. L. Mackey, M. Kjaer, The breaking and making of healthy adult human skeletal muscle in vivo. Skelet Muscle 7, 24 (2017).

29. A. L. Mackey, M. Kjaer, Connective tissue regeneration in skeletal muscle after eccentric contraction-induced injury. J Appl Physiol *(1985)* 122, 533–540 (2017).

30. A. L. Mackey, M. Magnan, B. Chazaud, M. Kjaer, Human skeletal muscle fibroblasts stimulate in vitro myogenesis and in vivo muscle regeneration. J Physiol 595, 5115–5127 (2017).

31. A. L. Mackey et al., Activation of satellite cells and the regeneration of human skeletal muscle are expedited by ingestion of nonsteroidal anti-inflammatory medication. FASEB J 30, 2266–2281 (2016).

32. A. Karlsen et al., Preserved capacity for satellite cell proliferation, regeneration, and hypertrophy in the skeletal muscle of healthy elderly men. FASEB J 34, 6418–6436 (2020).

33. R. M. Crameri et al., Myofibre damage in human skeletal muscle: effects of electrical stimulation versus voluntary contraction. J Physiol 583, 365–380 (2007).

34. R. M. Crameri et al., Changes in satellite cells in human skeletal muscle after a single bout of high intensity exercise. J Physiol 558, 333–340 (2004).

35. M. Saclier et al., Differentially activated macrophages orchestrate myogenic precursor cell fate during human skeletal muscle regeneration. Stem cells 31, 384–396 (2013).

36. M. Ahmadi et al., Aging is associated with an altered macrophage response during human skeletal muscle regeneration. Exp Gerontol 169, 111974 (2022).

37. S. Ancel et al., A dual-color PAX7 and MYF5 in vivo reporter to investigate muscle stem cell heterogeneity in regeneration and aging. Stem Cell Reports 19, 1024–1040 (2024).

38. S. Schiaffino, A. C. Rossi, V. Smerdu, L. A. Leinwand, C. Reggiani, Developmental myosins: expression patterns and functional significance. Skelet Muscle 5, 22 (2015).

39. J. N. Hathcock, Vitamin and Mineral Safety. D. M. A. W. H. Nguyen, Ed., (Council for Responsible Nutrition (CRN), ed. 3rd edition, 2024).

40. X. Ye, J. E. Maras, P. J. Bakun, K. L. Tucker, Dietary intake of vitamin B-6, plasma pyridoxal 5’-phosphate, and homocysteine in Puerto Rican adults. J Am Diet Assoc 110, 1660–1668 (2010).

41. M. S. Morris, M. F. Picciano, P. F. Jacques, J. Selhub, Plasma pyridoxal 5’-phosphate in the US population: the National Health and Nutrition Examination Survey, 2003-2004. Am J Clin Nutr 87, 1446–1454 (2008).

42. F. Hadtstein, M. Vrolijk, Vitamin B-6-Induced Neuropathy: Exploring the Mechanisms of Pyridoxine Toxicity. Adv Nutr 12, 1911–1929 (2021).

43. C. Canto, NAD(+) Precursors: A Questionable Redundancy. Metabolites 12, (2022).

44. L. Liu et al., Quantitative Analysis of NAD Synthesis-Breakdown Fluxes. Cell Metab 27, 1067–1080 e1065 (2018).

45. 45. J. B. Jensen, et al., A randomized placebo-controlled trial of nicotinamide riboside and pterostilbene supplementation in experimental muscle injury in elderly individuals. *JCI Insight* 7, (2022).

46. H. A. K. Lapatto et al., Nicotinamide riboside improves muscle mitochondrial biogenesis, satellite cell differentiation, and gut microbiota in a twin study. Sci Adv 9, eadd5163 (2023).

47. I. Sinha-Hikim, M. Cornford, H. Gaytan, M. L. Lee, S. Bhasin, Effects of testosterone supplementation on skeletal muscle fiber hypertrophy and satellite cells in community-dwelling older men. J Clin Endocrinol Metab 91, 3024–3033 (2006).

48. I. Sinha-Hikim et al., Testosterone-induced increase in muscle size in healthy young men is associated with muscle fiber hypertrophy. Am J Physiol Endocrinol Metab 283, E154–164 (2002).

49. R. W. Morton et al., A systematic review, meta-analysis and meta-regression of the effect of protein supplementation on resistance training-induced gains in muscle mass and strength in healthy adults. Br J Sports Med 52, 376–384 (2018).

50. T. M. Doering, P. R. Reaburn, S. M. Phillips, D. G. Jenkins, Postexercise Dietary Protein Strategies to Maximize Skeletal Muscle Repair and Remodeling in Masters Endurance Athletes: A Review. Int J Sport Nutr Exerc Metab 26, 168–178 (2016).

51. J. E. Tang, S. M. Phillips, Maximizing muscle protein anabolism: the role of protein quality. Curr Opin Clin Nutr Metab Care 12, 66–71 (2009).

52. O. C. Witard et al., Myofibrillar muscle protein synthesis rates subsequent to a meal in response to increasing doses of whey protein at rest and after resistance exercise. Am J Clin Nutr 99, 86–95 (2014).

53. J. Farup et al., Whey protein supplementation accelerates satellite cell proliferation during recovery from eccentric exercise. Amino Acids 46, 2503–2516 (2014).

54. S. Olsen et al., Creatine supplementation augments the increase in satellite cell and myonuclei number in human skeletal muscle induced by strength training. J Physiol 573, 525–534 (2006).

55. E. Marzetti et al., Restoring Mitochondrial Function and Muscle Satellite Cell Signaling: Remedies against Age-Related Sarcopenia. Biomolecules 14, (2024).

56. P. Sousa-Victor, L. Garcia-Prat, P. Munoz-Canoves, Control of satellite cell function in muscle regeneration and its disruption in ageing. Nat Rev Mol Cell Biol 23, 204–226 (2022).

57. P. Sousa-Victor, P. Munoz-Canoves, Regenerative decline of stem cells in sarcopenia. Mol Aspects Med 50, 109–117 (2016).

58. C. Suetta et al., Ageing is associated with diminished muscle re-growth and myogenic precursor cell expansion early after immobility-induced atrophy in human skeletal muscle. J Physiol 591, 3789–3804 (2013).

59. K. Fearon, J. Arends, V. Baracos, Understanding the mechanisms and treatment options in cancer cachexia. Nat Rev Clin Oncol 10, 90–99 (2013).

60. J. Brzeszczynska et al., Loss of oxidative defense and potential blockade of satellite cell maturation in the skeletal muscle of patients with cancer but not in the healthy elderly. Aging (Albany NY) 8, 1690–1702 (2016).

61. M. Beltra et al., NAD(+) repletion with niacin counteracts cancer cachexia. Nat Commun 14, 1849 (2023).

62. J. M. Park, Y. M. Han, H. J. Lee, Y. J. Park, K. B. Hahm, Nicotinamide Riboside Vitamin B3 Mitigated C26 Adenocarcinoma-Induced Cancer Cachexia. Front Pharmacol 12, 665493 (2021).

63. R. J. Butterfield, Congenital Muscular Dystrophy and Congenital Myopathy. Continuum (Minneap Minn) 25, 1640–1661 (2019).

64. M. Ganassi, F. Muntoni, P. S. Zammit, Defining and identifying satellite cell-opathies within muscular dystrophies and myopathies. Exp Cell Res 411, 112906 (2022).

65. A. Sacco et al., Short telomeres and stem cell exhaustion model Duchenne muscular dystrophy in mdx/mTR mice. Cell 143, 1059–1071 (2010).

66. N. A. Dumont, M. A. Rudnicki, Targeting muscle stem cell intrinsic defects to treat Duchenne muscular dystrophy. NPJ Regen Med 1, 16006-(2016).

67. K. Kodippili, M. A. Rudnicki, Satellite cell contribution to disease pathology in Duchenne muscular dystrophy. Front Physiol 14, 1180980 (2023).

68. N. M. Vieira et al., Jagged 1 Rescues the Duchenne Muscular Dystrophy Phenotype. Cell 163, 1204–1213 (2015).

69. N. Hayashiji et al., G-CSF supports long-term muscle regeneration in mouse models of muscular dystrophy. Nat Commun 6, 6745 (2015).

70. J. von Maltzahn, J. M. Renaud, G. Parise, M. A. Rudnicki, Wnt7a treatment ameliorates muscular dystrophy. Proc Natl Acad Sci U S A 109, 20614–20619 (2012).

71. M. Loreti, A. Sacco, The jam session between muscle stem cells and the extracellular matrix in the tissue microenvironment. NPJ Regen Med 7, 16 (2022).

72. C. S. Fry, T. J. Kirby, K. Kosmac, J. J. McCarthy, C. A. Peterson, Myogenic Progenitor Cells Control Extracellular Matrix Production by Fibroblasts during Skeletal Muscle Hypertrophy. Cell Stem Cell 20, 56–69 (2017).

73. O. Midttun, S. Hustad, P. M. Ueland, Quantitative profiling of biomarkers related to B-vitamin status, tryptophan metabolism and inflammation in human plasma by liquid chromatography/tandem mass spectrometry. Rapid Commun Mass Spectrom 23, 1371–1379 (2009).

74. J. Bergstrom, Percutaneous needle biopsy of skeletal muscle in physiological and clinical research. Scand J Clin Lab Invest 35, 609–616 (1975).

