## Supplemental figures for "Nicotinamide and Pyridoxine supplementation stimulates muscle stem cells in a randomized clinical trial on muscle repair"

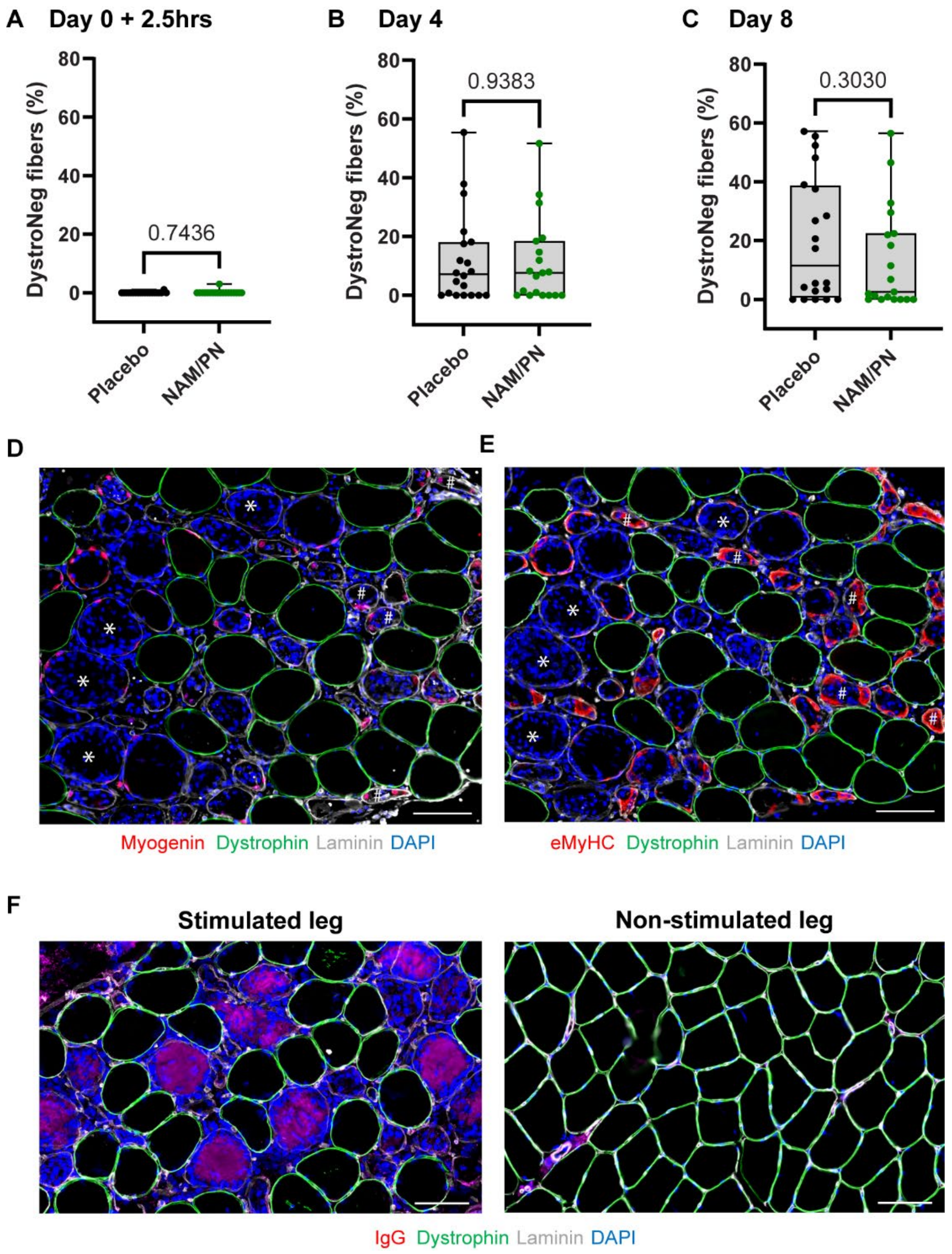


**Figure S1: The number of Dystrophin-negative fibers is not affected by NAM/PN treatment.** (A-C) The percentages of damaged fibers as assessed by the proportion of dystrophin-negative (DystroNeg) fibers on cross sections of muscle biopsies collected at (A) Day 0 + 2.5hrs, (B) Day 4 and (C) Day 8 (stimulated leg) were quantified and compared between NAM/PN vs. placebo group. Individual dots represent individual subjects and data are expressed as the percentage of dystrophin-negative fibers out of total fibers. Placebo group, n=20; NAM/PN group, n=19. (D,E) Representative images illustrating the massive infiltration of nuclei inside dystrophin-negative fibers undergoing inflammatory infiltration and necrosis (examples shown by *), and the overlap with active regeneration demonstrated marked by myogenin$^{+}$ nuclei and eMyHC-positive fibers (examples shown by #). (F) Necrotic fibers were assessed by the presence of IgG-positive fibers on cross sections of muscle biopsies collected at Day 8 (stimulated and non-stimulated legs). Scale bars = 100µm.

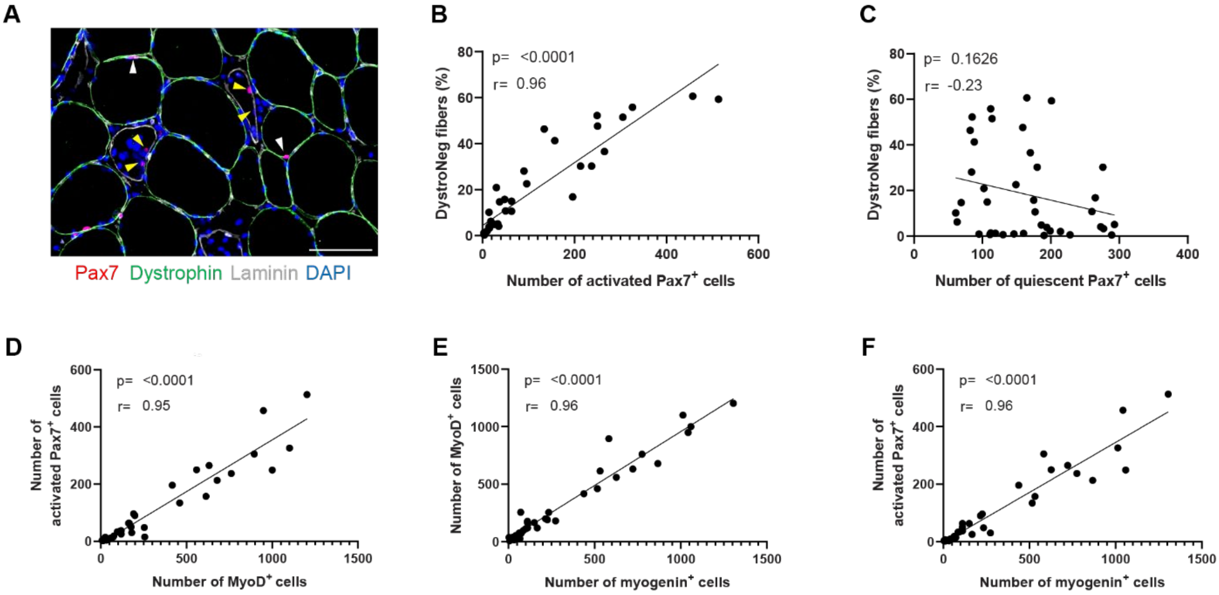


**Figure S2: the muscle injury protocol triggers myogenic activation and differentiation of MuSCs** (A) Quiescent vs. activated Pax7$^+$ cells were distinguished as mononucleated cells belonging to dystrophin-positive fibers and residing under the basal lamina (indicated by white arrowheads) whereas activated Pax7$^+$ cells were distinguished as being located inside or in close proximity of dystrophin-negative fibers (indicated by yellow arrowheads). (B-F) Correlation analyses (placebo group, n=20; NAM/PN group, n=19) between the percentage of dystrophin-negative fibers and the number of (B)activated Pax7$^+$ cells , (C) quiescent Pax7$^+$ cells; between the number of activated Pax7$^+$ cells and the number of (D) MyoD$^+$ cells and (E) myogenin$^+$ cells; and between (F) the number of MyoD$^+$ cells and the number of myogenin$^+$ cells. Scale bars = 100µm.

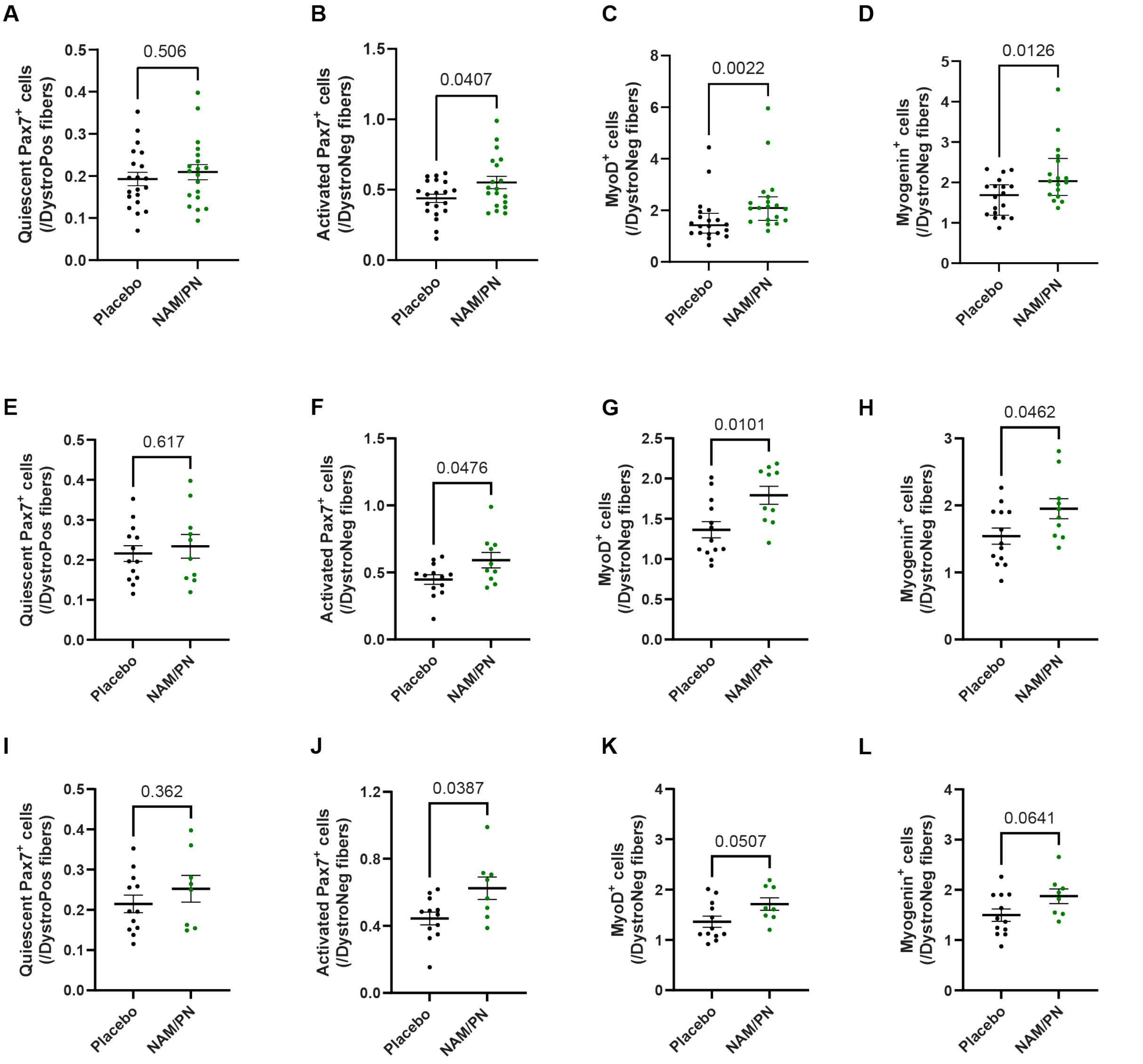


**Figure S3: NAM/PN supplementation increases the number of activated Pax7$^+$ cells as well as the number of MyoD$^+$ and myogenin$^+$ cells.** The number of MuSCs and their progenies were quantified by immunohistofluorescence on cross section of biopsies collected at Day 8 (stimulated leg). Analyses were performed on three different sets of subjects: (A-D) all subjects from per-protocol population (placebo group, n=20; NAM/PN group, n=19); (E-H) subjects with 5% or more of dystrophin-negative fibers (placebo group, n=13; NAM/PN group, n=10); (I-J) subjects at or above the median value for the percentage of dystrophin-negative fibers (placebo group, n=12; NAM/PN group, n=8). Comparison between NAM/PN vs. placebo group of the number of (A, E, I) quiescent Pax7$^+$ cells, (B, F, J) activated Pax7$^+$ cells, (C, G, K) MyoD$^+$ cells, and (D, H, L) myogenin$^+$ cells. Data are represented as (A,B, E-L) mean ± SEM or (C,D) median with interquartile range, with individual dots representing individual participants and are expressed relative to the number of (A, E, I) dystrophin-positive (DystroPos) and to the number of (B-D; F-H, J-L) dystrophin-negative (DystroNeg) fibers.

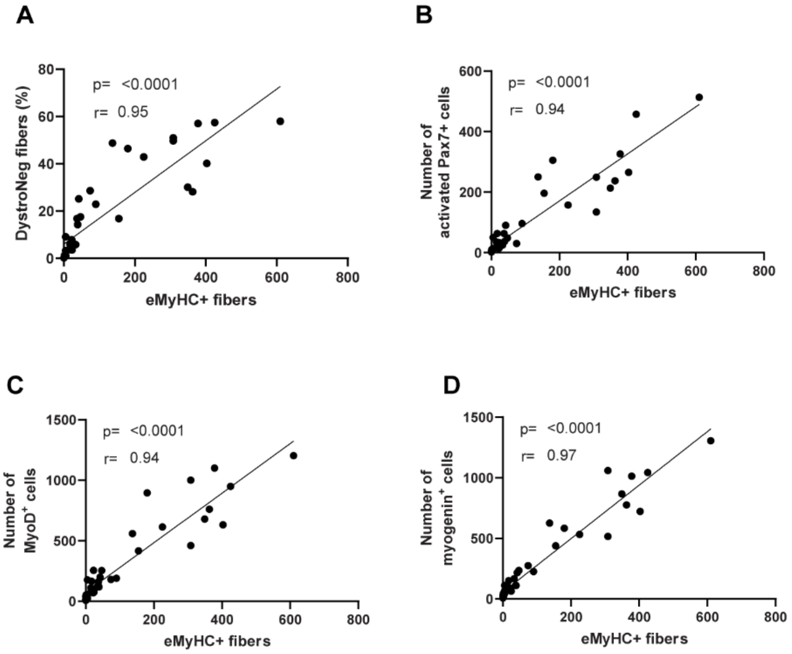


**Figure S4: Evaluation of fiber regeneration by immunohistofluorescence on cross sections of biopsies collected at Day 8 (stimulated leg).** (A) Correlation analysis between the percentage of dystrophin-negative fibers and the number of eMyHC$^+$ fibers. (B-D) Correlation analyses between the number of eMyHC$^+$ fibers and the number of (B) activated Pax7$^+$ cells, (C) MyoD$^+$ cells and (D) myogenin$^+$ cells. Individual dots represent individual participants. Placebo group, n=20; NAM/PN group, n=19.
